# Augmenting gait in a population exhibiting foot drop with adaptive functional electrical stimulation

**DOI:** 10.1101/2022.04.27.22273623

**Authors:** Jeremiah Robison, Ren Gibbons, Dean Achelis, Brinnae Bent, Doug Wajda, Rebecca Webster

## Abstract

Functional electrical stimulation (FES) can be used for improvement of gait in people with foot drop resulting from neurological disease or injury. However, existing FES solutions suffer from significant limitations. First, fixed placement electrode devices are difficult to tune stimulation for improved ankle dorsiflexion while maintaining balanced ankle eversion. Second, standard tilt-sensor-like triggers do not allow for adaptive, configurable stimulation sequencing algorithms.

Here, we introduce the first FES system with adaptive current steering for an array of FES electrodes enabling precise control over dorsiflexor and evertor muscles, allowing for personalized treatment to correct key foot drop characteristics including dorsiflexion at heel strike and ankle inversion during swing phase.

We share results of a pre-test, post-test study on thirty-two participants exhibiting symptoms of foot drop. The differences in pre-test versus post-test primary and secondary outcome measures were statistically significant (p<0.0125) within our cohort. With adaptive, current-steering FES, ankle dorsiflexion at heel strike increased an average 5.2°, and ankle inversion during swing phase was reduced by an average -3.6°, bringing the ankle to a more neutral position for stabilization.

By significantly increasing ankle dorsiflexion at heel strike and decreasing ankle inversion during swing phase, adaptive FES enabled a more neutral ankle at heel strike, which is associated with greater ankle stability and decreased fall risk. Gait augmentation using adaptive, current-steering FES improved gait in a population exhibiting symptoms of foot drop.

## I. Introduction

Neurological disease or injury can significantly impact gait through reduced voluntary movement or spasticity, which can impede physical safety and reduce the ability to participate in activities of daily living, which can hinder community engagement and emotional well-being.

Foot drop describes difficulty lifting the front part of the foot and is commonly seen in adults with muscle weakness related to upper motor neuron disease or injury, including stroke survivors and individuals with multiple sclerosis[1,2]. Specifically, foot drop negatively impacts gait through decreased voluntary ankle dorsiflexion and excessive ankle inversion[3]. Decreased dorsiflexion can result in lessened dynamic loading response[4] and increased fall risk[5]. Excessive inversion can lead to lateral instabilities, stretching of lateral ankle structures, ankle sprains, and ultimately pain during weight acceptance[6].

Foot drop severity can be quantitatively assessed with several kinematic metrics. The primary outcome metrics for this study are dorsiflexion at heel strike[7] and mean inversion during swing phase[8]. The secondary outcome metrics are foot angle at heel strike and heel-toe time (single-side heel strike to toe strike time). Foot angle at heel strike has study relevance since lower foot angles can indicate decreased toe clearance during the swing phase of gait[9]. Similarly, heel rocker is a gait function that improves the fluidity of weight acceptance[10], which is often diminished in individuals with foot drop. Heel-toe time is a method for approximating heel rocker function.

Various assistive technologies exist to address gait deviations in individuals with foot drop. These include orthoses such as AFOs, assistive devices such as canes and walkers, and functional electrical stimulation (FES) devices[11]. FES devices improve gait by applying low amplitude electrical pulses to skeletal muscles to induce involuntary contraction to replace interrupted pathways from the central nervous system to peripheral neurons. Contrary to passive devices, FES provides dynamic stabilization and enables more normative movement of the ankle during gait[12].

FES gait augmentation systems offer numerous benefits over passive orthoses to individuals with impacted mobility[11]. FES gait training has been shown to improve walking speed and duration compared to traditional orthotic gait devices[13]. FES has been linked to increased endurance[14, 15], effective neural retraining[16], and increased bone density[17, 18]. Despite these benefits, currently available FES systems have significant limitations: stimulation tuning for improved ankle dorsiflexion while maintaining balanced ankle eversion is challenging for devices with a single pair of manually placed gel electrodes[19]. Additionally, other FES devices lack adaptive personalized algorithms for fine-tuned temporal stimulation sequencing[20].

The Cionic Neural Sleeve is the first FES system that provides adaptive, multi-channel current-steering FES to an array of configurable electrodes to enable precise control of the dorsiflexors and evertors (including the tibialis anterior and the peroneus longus). The algorithms controlling temporal stimulation sequencing are also personalizable and adapt seamlessly across gait speeds. This enables the system to reduce key foot drop characteristics, including ankle dorsiflexion and inversion, and personalize treatment for foot drop.

The primary aim of this study was to investigate whether adaptive, current-steering FES of the lower leg improves gait in participants with foot drop, as measured by ankle dorsiflexion, ankle inversion, foot angle, and heel-toe time.

## II. METHODS

### A. Participant Recruitment

A total of 60 participants were assessed for eligibility. Study inclusion criteria specified participants must be an adult aged 18-70 living with a lower extremity impairment that makes walking difficult or uncomfortable; be capable of sitting, standing, and walking independently or with assistance; be able to walk at least 50 feet independently or with assistance; and be able to understand and follow basic instructions in English. Of those assessed, eight were excluded due to not meeting eligibility requirements, declining to participate, or other. Fifty-two were enrolled in the study. Of the enrollees, fourteen were lost to follow up due to scheduling logistics. Thirty-eight were allocated for data collection. Four of the allocated were excluded for insufficient endurance to complete the experimental protocol, and two were excluded due to equipment errors. The final analysis pool contained 32 subjects (Fig. 1).

**Fig. 1.**
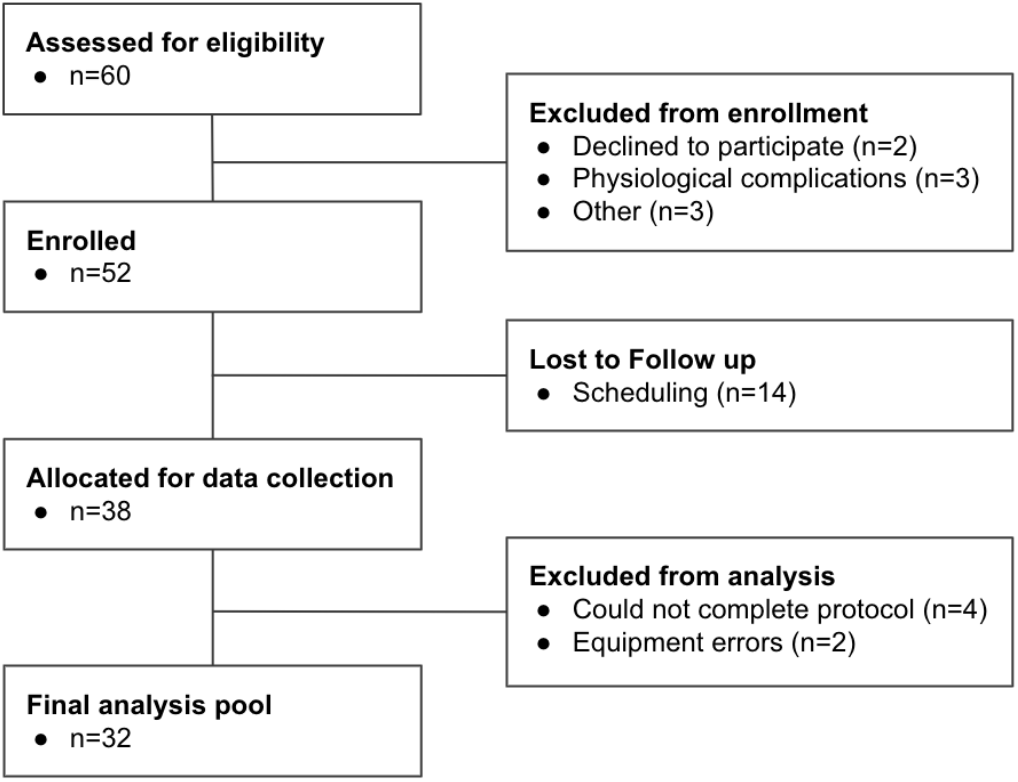
Flow chart on participant inclusion. Of 60 assessed for eligibility, 52 were enrolled. Thirty-eight of the enrollees were allocated for data collection, and 32 were included in the final pool. Reasons for exclusion from analysis include inability to complete protocol and equipment errors.

### B. Ethical Considerations

The present study was carried out according to the Declaration of Helsinki. Participation was voluntary and based on written informed consent from the participant. The study has been approved by the Institutional Review Board at Ethical & Independent Review Services (E&I 21039-01A) and the Institutional Review Board at Cleveland State University (IRB-FY2021-301).

### C. Study Design

The study design is a one-group, pre-test post-test study where we assessed the impact of FES relative to each participant’s unstimulated gait in a pairwise fashion. All participants received adaptive FES to the dorsiflexors and evertors during up to three back-to-back walking sets.

Prior to walking with FES, each participant underwent a standard seated FES configuration protocol facilitated by trained researchers to set stimulation parameters to achieve observable dorsiflexion (ideally exceeding 10°) with balanced eversion while maintaining participant comfort. Frequency and pulse width parameters were set to 40 Hz and 300 μs, respectively, and intensity was increased until the desired ankle movement was achieved. Frequency and pulse width could also be tuned in increments of 5 Hz and 100 μs as needed for comfort or efficacy. FES configuration was completed using the Cionic mobile application. Each participant gained familiarity with stimulated gait by traversing a 40-foot walkway as a warmup. They walked with their typical assistive devices, if applicable, excluding AFOs.

Next, each participant alternated between unstimulated (noFES) walking collection of their typical gait and stimulated (FES) walking collection. Data was collected from the Cionic Neural Sleeve using the following sensors: Bosch Sensortec inertial measurement units (IMUs) placed on the thigh, shank, and foot to reconstruct the leg’s spatial orientation and three foot pressure sensor pads sampled using an analog-to-digital converter (ADS1298CZXGT, Texas Instruments) in the shoe for gait event detection. IMUs were calibrated prior to each participant’s collections. For the stimulated collections, the dorsiflexors were stimulated to maintain toe clearance during swing phase and improve heel strike, and evertors to achieve neutral ankle at heel strike and loading response. Unstimulated and stimulated collections were performed back-to-back up to three times (six total collections) to directly quantify the impact of FES. Each collection consisted of walking across the walkway, turning around, and walking back to the starting point. Time intervals corresponding to the turnaround were identified via manual inspection of kinematics data and were removed from the analysis. The first two steps and last step for each length of the walkway were removed from the analysis. Pressure sensor data was automatically labeled and manually inspected to extract heel strike, toe strike, and toe off events.

### D. Adaptive Functional Electrical Stimulation

In this study, we use the Cionic Neural Sleeve, an adaptive, current-steering FES device to stimulate dorsiflexors and evertors to improve ankle movement. The novel technology includes an array of FES gel electrodes placed on the anterolateral region of the impaired lower leg (Fig. 2), corresponding with the atlas of the muscle motor points for the lower limb[21]. The frequency (Hz) and pulse width (μs) of all electrodes are tunable. Additionally, the current intensity (mA) of each electrode in the array can be independently configured using a mobile application. This multi-channel current-steering allows for precision control of ankle movement in multiple planes. The sequencing of muscle stimulation is controlled by algorithms operating on data streamed from on-body sensors. Specifically, the algorithms use spatial orientation data to trigger muscle contraction and relaxation events that adapt to changes in gait speed.

**Fig. 2.**
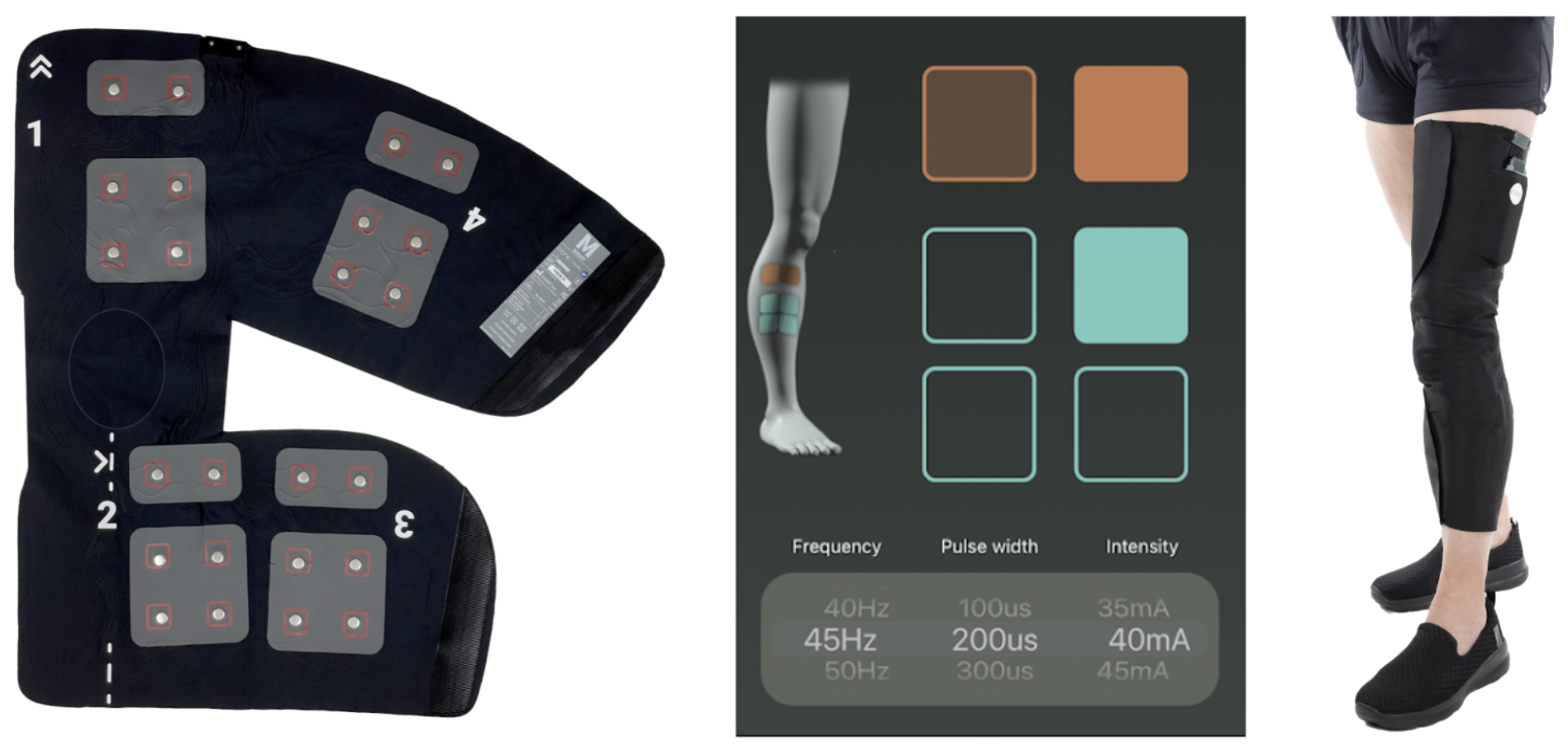
Left: Cionic Neural Sleeve showing the configuration of the FES and EMG electrode arrays. Center: Cionic mobile application interface for setting FES parameters (frequency, pulsewidth, intensity) and current field steering. Right: Cionic Neural Sleeve on body.

### E. Statistical Analysis

Based on our statistical power analysis, we required eighteen or more participants to achieve 80% power to reject the null hypothesis which states there is no difference between pre-test (noFES) and post-test (FES) conditions in a pairwise comparison (α = 0.05). We selected an effect size of 0.7 for the power analysis based on a previous study of FES impact on post-stroke participants[22].

In this analysis, we examined whether the impact of FES on gait is statistically significant relative to unstimulated gait by pairwise comparison. For each metric, we averaged each participant’s metric values across steps for both noFES and FES walking. Each metric’s final analysis pool contained 32 noFES and 32 FES samples. The Shapiro-Wilk test for normality[23] was used to assess the assumption of normality and paired t-tests were used to statistically compare noFES and FES foot drop metrics if the assumption for normality was met (all metrics except foot angle at heel strike). If the normality assumption was not met, the Wilcoxon signed rank test was used as an alternative to the paired t-test (this was the case for foot angle at heel strike). Bonferroni multiple hypothesis correction was performed (four metrics tested), and our corrected significance level for comparison was 0.0125 (0.05 divided by the four hypotheses tested). We also report the 95% confidence interval for the differences between FES and noFES foot drop metrics.

## III. RESULTS

Current-steering FES enables precise control over muscles to personalize the balance between dorsiflexion and inversion. The primary outcomes of this study were dorsiflexion at heel strike and inversion during swing phase, as these metrics are directly related to fall risk and ankle stabilization, respectively. Our secondary outcomes of this study include foot angle at heel strike and heel-toe time which also correlate to gait stability, dynamic loading response, and improved rocker[23].

### A. Participant Demographics

The final analysis pool was split across 32 participants aged 61.1±10.9 living near San Francisco and Cleveland. The subjects included 18 females and 14 males. All subjects had a diagnosis of neurological disease or injury: 20 were post-stroke, eight had multiple sclerosis, two had a spinal cord injury, one had cerebral palsy, and one had idiopathic foot drop. All exhibited symptoms of foot drop. The average stimulation parameters across participants included an amplitude of 44±11 mA, frequency of 38±14 Hz, and a pulse duration of 306±51 μs.

### B. Primary Outcomes

The impacts of FES on dorsiflexion across 32 participants exhibiting symptoms of foot drop are shown in Fig. 3A. The kinematics profile shows average dorsiflexion throughout the gait cycle for all participants, and the shaded region shows the standard deviation. Zero percent corresponds to heel strike. Additionally, the participants’ baseline noFES kinematics showing symptoms of foot drop are in orange while Cionic FES show the impact of gait assist in blue. We observed that the primary impact of FES on dorsiflexion occurs at heel strike and during swing phase (approximately to 65-100% of gait), and that dorsiflexion at heel strike is increased by an average of 5.2° (Confidence Interval (CI)[3.5°, 6.8°]) with statistical significance (p-value=7e-5). The average increase of 5.2° is greater than the minimal clinically important difference of 4.9° as defined by the Clinical Practice Guideline for the Use of Ankle-Foot Orthoses and Functional Electrical Stimulation Post Stroke[24]. 93.8% of participants showed increased dorsiflexion at heel strike; the biggest observed increase from a participant was an average of 14.6°.

**Fig. 3.**
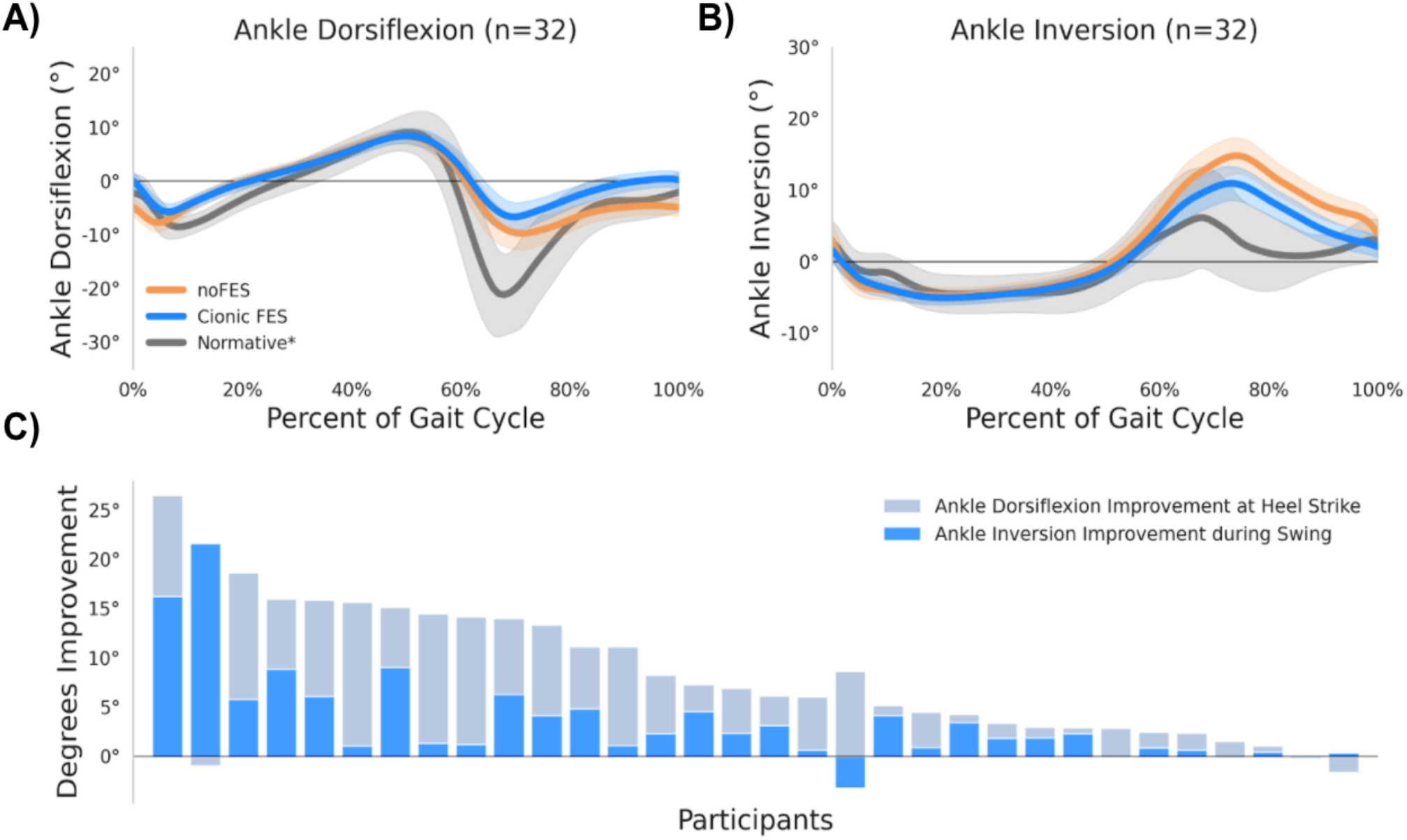
(A) Ankle dorsiflexion gait profiles averaged over the 32 participants walking with Cionic FES (blue profile) and without (orange profile). The gait cycle profile is represented as a percent, where 0% occurs at heel strike. The solid lines represent the cohort average, and the shaded intervals are ± one standard deviation. (B) Ankle inversion gait profiles averaged over the 32 participants with Cionic FES (blue) and without (orange). (C) Stacked degrees of dorsiflexion improvement (gray bar) and inversion improvement (blue bar) for each participant. All participants but one showed a net improvement across both primary outcomes. *The gray normative profile was determined from the typical walking gait of eleven normative participants. This was a sample of convenience conducted on Cionic employees with no known gait impairments. Normative participant data was collected under IRB oversight and consisted of 11 participants (10 male, 1 female) with age range [24-53].

In addition to decreased dorsiflexion, the typical participant also exhibits excessive swing phase ankle inversion as shown in Fig. 3B. Evertor FES is configured to reduce ankle inversion during swing phase through loading response (Fig. 3B) as this increases lateral loading response stability. We show an improved outcome by reducing mean inversion during the swing phase by an average of -3.6° (CI[-5.3°, -1.8°], p-value=3e-4). The largest observed decrease was an average of -21.6°. In total 88% of participants showed a positive improvement to ankle inversion during swing.

Fig. 3C shows the stacked degrees of improvement for dorsiflexion and inversion for each participant, providing a more comprehensive picture of the impact of gait assist across multiple movements. Some participants exhibit large improvements to inversion due to high baseline inversion, while others show higher dorsiflexion improvement relative to inversion. 96.9% showed net improvement across both primary metric outcomes (net average 9° improvement).

### C. Secondary Outcomes

We observed an average of 5.5° (CI[3.9°, 7.0°]) increase in foot angle at heel strike (p-value=9e-7). Heel-toe time increased by an average of 3.4% of gait (CI[1.9%, 4.8%], p-value=4e-5). After FES treatment, average foot angle at heel strike and heel-toe time were closer to the normative average. With dorsiflexor stimulation, foot angle increased in 96.9% of participants. The largest FES-driven increase in foot angle was 15.8°. Additionally, stimulated gait resulted in increased heel-toe time in 90.6% of participants. The participant with the greatest increase in heel-toe time showed a 17.5% increase.

## IV. DISCUSSION

This study aimed to evaluate the effects of a first-of-its-kind adaptive, multi-channel current-steering FES on gait in a population exhibiting foot drop. We demonstrated statistically significant improved gait in 32 participants exhibiting symptoms of foot drop due to neurological disorders. On average, these participants showed a 5.2° increase in dorsiflexion at heel strike and a 3.6° reduction in ankle inversion during swing phase. Together, this brings the ankle into a more stable, neutral position, which is associated with decreased fall risk[5]. The increase in dorsiflexion also resulted in a 5.5° increase in foot angle at heel strike which correlates to better foot clearance, and an increase in heel-toe time of 3.4% of the gait cycle has links to more dynamic loading response.

Neurological disease or injury can significantly impact gait, impeding physical safety and reducing the ability to participate in activities of daily living, which hinders community engagement and emotional well-being. Foot drop, a major inhibitor of mobility and a contributor to fall risk, is commonly seen in people with multiple sclerosis, post stroke, or other neurological conditions. Existing solutions for gait correction of foot drop include passive devices and FES systems, which stimulate skeletal muscles to restore motor function[11]. FES provides dynamic stabilization, enabling movement while improving stability. While FES systems show enormous potential for gait improvements[24, 25], current marketed products have limited capabilities for real-life mobility enhancements[19, 20]. Given the large number of people living with mobility impairments and the significant accessibility challenges and financial burdens they face[26], it is critically important to find solutions that address the shortcomings of current assistive gait devices.

The Cionic Neural Sleeve FES system used in this study utilizes adaptive current-steering to enable precise control over the dorsiflexors and evertors. Current-steering allows for personalization of the balance between increased dorsiflexion and reduced ankle inversion, leading to a more neutral ankle position at heel strike in order to reduce fall risk. While many FES solutions that enable personalized control of stimulation are available only in the clinical setting, the Cionic Neural Sleeve is available for at-home use, extending the applications of FES to include improved daily mobility and remote rehabilitation.

### A. Study Limitations

This study did not include the collection of certain traditional gait analysis metrics including spatial measures like gait speed and stride length as we relied solely on the on-body device for all data collection and metric extraction. Additionally, we were not able to compare FES treatment impacts relative to standard of care, i.e., AFO, in a randomized control trial. A randomized controlled trial remains a future research study.

## V. CONCLUSION

The ability to reduce foot drop, a major inhibitor of mobility and a contributor to fall risk, would provide great value to those with impaired gait. This is the first study evaluating the effects of adaptive, current-steering stimulation from the Cionic Neural Sleeve on gait in patients exhibiting symptoms of foot drop. Precise control over the dorsiflexor and evertor muscles results in statistically significant improvements to ankle movement, including increased dorsiflexion and decreased inversion, which can stabilize the ankle and decrease fall risk.

## Data Availability

All data produced in the present study are available upon reasonable request to the authors.

## Notes

This work was supported by CIONIC, Inc.

### Competing Interest Statement

J.R., and R.W. are employed by CIONIC Inc. R.G. and B.B. are consultants for CIONIC Inc. D.A. is a former employee of CIONIC Inc. D.W. research is funded in part by CIONIC Inc.

### Clinical Trial

NCT05346640

### Funding Statement

This study was funded by CIONIC, Inc.

### Author Declarations

The present study was carried out according to the Declaration of Helsinki. Participation was voluntary and based on written informed consent from the participant. The study has been approved by the Institutional Review Board at Ethical &; Independent Review Services (E&I 21039-01A) and the Institutional Review Board at Cleveland State University (IRB-FY2021-301).

